# Heart Block Identification from 12-Lead ECG: Exploring the Generalizability of Self-Supervised AI

**DOI:** 10.1101/2024.10.15.24315546

**Authors:** Teya S. Bergamaschi, Collin M. Stultz, Ridwan Alam

## Abstract

Timely diagnosis and treatment of heart blocks are critical for preventing fatal outcomes in patients with cardiac conduction disorders. Expert analysis of clinical 12-lead electrocardiograms (ECG) remains the standard diagnosis apparatus. In this study, we propose a self-supervised deep learning model that can detect evidence of heart blocks in ECG signals. We build a multichannel residual neural network (KRes) and train this model in a self-supervised contrastive fashion leveraging 3.6 million ECG from patients at the Massachusetts General Hospital (MGH) to learn robust ECG representations. We evaluate the utility of such representations on a large public dataset, PTB-XL, through fine-tuning and linear probing for identifying heart blocks. To compare, we also build a baseline supervised model by adapting KRes. The training data contains a subset of 10.6 thousand PTB-XL ECG from patients with and without heart block patterns, and a holdout set of 1319 ECG is used for evaluation. We compare the performances of the finetuning and the linear probing of the self-supervised representations with that of the supervised model using the area under the receiver-operating curve (AUC), sensitivity, specificity, and predictive values, and found those performing equally well. Moreover, we repeat the training cycles of the three pipelines while reducing the number of training samples and demonstrate that the performance of the self-supervised representations remain steady even when labeled data is limited. This research underscores the potential of self-supervised learning in cardiac diagnostics, emphasizing generalizability and performance across diverse datasets and clinical settings, especially in data-scarce paradigms.

## I. Introduction

Action potentials, spontaneously initiated at the sinoatrial node of the heart, traverse the heart chambers via the cardiac conduction pathway. This electric propagation triggers and regulates the contractions of the atria and the ventricles, keeping the heart pumping. Blockage along the conduction pathway, known as heart block or conduction disorder (CD), impedes the rhythm and order of contractions, causing irregular heartbeats, arrhythmia, and can lead to syncope, cardiac arrest, and stroke [1]. Depending on the location of the blockage, the symptoms, treatments, and severity vary. For example, 1^st^ degree atrioventricular (AV) block is prevalent in 6% population older than 60 years, though often not severe. However, a 3^rd^ degree aka complete AV block (prevalence 0.03%) can collapse the ventricular contraction, leading to sudden cardiac arrest -a life-threatening condition [2].

A 12-lead electrocardiogram (ECG) captures the changes in the cumulative electrical dipole across the heart with time and is used as the standard apparatus to diagnose various CD and other arrhythmias [3]. Cardiologists manually analyze the ECG by identifying potential signs of blockage in the ECG and suggest treatments accordingly. The sign of 1^st^ degree AV block is a PR interval greater than 200 ms on the ECG without disruption of atrial to ventricular conduction. State-of-the-art automated diagnosis algorithms attempt to extract similar characteristic patterns from the ECG by detecting fiducial points and calculating the relevant intervals such as PR, QT, QRS duration, etc. [4] While such algorithms can provide clinically meaningful interpretations, they fail to generalize against variations across patients, symptoms, institutions, and signal quality, hence mandating clinician oversight of such machine-generated diagnostic reads [5].

Recent applications of deep learning methods on health records and clinical time series have also motivated many ECG models [6]–[9]. Many models have been proposed to identify “normal” ECG against multiple “abnormalities”, identifying arrhythmias, conduction disorders, among others. A major challenge for such models is the scarcity of data for any cardiac disease category in any dataset from a single clinical institute, as the corresponding prevalences vary significantly. If a model is trained on a dataset that has a large number of “normal” ECGs and a minuscule subset from patients with complete AV block, the performance achieved on validation sets with low pre-test probabilities struggle to translate to clinically useful post-test probabilities [10]. Moreover, the generalizability of such solutions to patients from other institutes or data from different sources remains a major challenge. As these methods depend heavily on the “training” dataset, they often suffer a drop in performances when applied on data from different sources or distributions.

In this work, we address these challenges by proposing a self-supervised method for identifying heart blocks from 12-lead ECG signals. First, we train a contrastive representation learning pipeline (Fig. 1) on a large dataset of 3.6 million ECGs, which was acquired from about 800 thousand patients at the Massachusetts General Hospital (MGH). Then, we apply that pipeline to get ECG representations from a publicly available ECG dataset, namely PTB-XL, which contains 21 thousand ECGs from 19 thousand patients along with their diagnostic labels, which includes patients with various conduction disorders (CD) as well as patients with normal ECGs [11]. To demonstrate the utility of these representations, we compare a simple classifier trained on those representations against a large supervised Resnet model trained on 12-lead ECGs. We also explore the potential benefits of “fine-tuning” the self-supervised pipeline for this classification task. The area under the receiver-operating curve (AUC), sensitivity, specificity, and positive and negative predictive values are calculated for a detailed evaluation. Finally, we demonstrate the generalizability of this approach by comparing the performance of the models across different training set sizes. Our work highlights the potential of deep learning-based 12-lead ECG representations as a performant and generalizable apparatus in cardiac diagnostic applications.

**Fig. 1.**
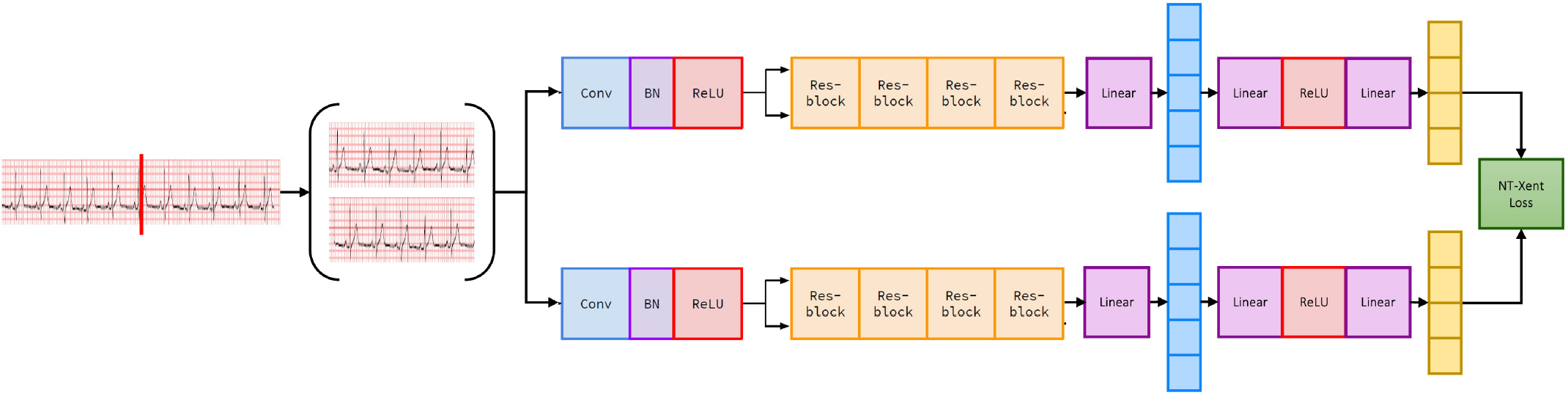
Architecture overview of self-supervised model.

## II. Related Works

Recent research on automated ECG analyses has been adopting deep learning solutions. Supervised models have been trained, where sufficient labels were available, for tasks ranging from identifying arrhythmias [7] to diagnosing structural defects [12] and estimating hemodynamic parameters [9], [13]. Early-stage convolutional neural networks, trained in a supervised manner, have been reported to achieve more than 0.98 AUC for multiple abnormality identification, even performing better than some clinicians [6], [7], [14]. To ensure a common standard, [8] has presented some benchmark results for such applications, though the generalizability of those models for other datasets remains unexplored. Moreover, many of these models evaluate their performance on a much smaller test-set; for example, [7] tested only on 328 patients while training on 54 thousand patients and [14] had a test-set of only 827 ECG samples.

The scarcity of clinically validated annotations, especially for rare cardiac conditions and diseases, poses a perpetual challenge for learning-based methods. Self-supervised learning has been investigated in different domains in bypassing the need for large labeled datasets. ECG representations learned this way have proven useful for downstream tasks, especially when finetuned for specific applications [15], [16]. The generalization of such representations to external datasets, as well as in label-deficient settings, still needs to be demonstrated.

## III. Methods

To analyze whether a 12-lead ECG signal contains evidence of cardiac conditions, we explore two learning paradigms: supervised and self-supervised models.

### A. Data Description

We use two datasets in this work: the PTB-XL dataset to train and validate our methods’ performances, and the MGH dataset to train the self-supervised representation learning pipeline.

PTB-XL [11] is a large ECG dataset, publicly available on Physionet, that contains 21,837 clinical 12-lead ECG from 18,885 patients with diagnoses of conduction disorders, myocardial infarctions, ischemia, and hypertrophic cardiomyopathy, as well as those without any cardiac disorders. For our experiments, we curate a subset of 13,240 ECGs whose labels are validated by at least one cardiologist with 100% confidence scores; including 4630 ECGs that are labeled as positive for one or more of the CD categories: fascicular blocks, bundle branch blocks, AV blocks, Wolff-Parkinson-White syndromes, or non-specific CD. And 8610 ECG were labeled as “normal” or without any cardiac disorders.

For the self-supervised representation learning, we utilized a much larger dataset, referred to as the MGH dataset, which contains 3,617,253 12-lead ECG recordings, from 770,615 patients at the Massachusetts General Hospital (MGH), acquired between January 1981 and December 2020. All machine-generated measurements were signed off by cardiologists, who often verified the measurements and interpretations if deemed necessary. Retrospective analyses of this dataset were approved by the Institutional Review Board (IRB) at the Mass General Brigham (protocol #2020P000132).

### B. Model Architecture

We present a deep learning architecture, KRes, a multichannel E<u>K</u>G <u>res</u>idual neural network (ResNet) consisting of four residual blocks, that we use for both our supervised and self-supervised learning pipelines. KRes is an updated version of our earlier work [9]. Each residual block in KRes contains two convolution layers and a skip connection. We add twelve 1-d convolution layers before the residual blocks to ingest the 12-lead ECG signal as a multichannel input. We use a kernel size of 16 units for all convolutional filters. The input channels are of 12×2500 sample length, as we resample all 10-second ECG signals to a 250 Hz sampling rate to get the input tensor. The ingest layer convolves this tensor with a single sample stride over 64 filters. We learn convolution layers with 128, 196, 256, and 320 filters for the four consecutive residual blocks. For each block, the skip connections are implemented with max pooling and a 1-to-1 convolution layer. Batch normalization and ReLU activation layers follow each convolution layer of the model. Using average pooling, we get a 1×320 feature representation from the output of the last residual block.

For identifying evidence of conduction disorder (CD) from 12-lead ECG signals, we adopt the KRes backbone in building a 12-lead ECG supervised classifier model. As presented above, the KRes backbone learns a 1×320 representation vector from the 12-lead ECG input. The representation from the residual blocks is passed on to two fully connected dense layers followed by a rectified linear unit (ReLU) layer that outputs the probability of the presence of CD in comparison to normal ECG and thus completes the end-to-end supervised classification architecture. This supervised model is used as the performance baseline for our proposed self-supervised model. As our core contribution, we implement a self-supervised architecture following the contrastive learning standard method, SimCLR [17]. We design to contrast two non-overlapping 5-second segments of the same 10-second ECG [Fig 1]. The goal of contrastive learning is to learn useful representations of the input data by maximizing agreement between embeddings from the same data. This process involves using a backbone to generate representations, a projection head to produce embeddings, and a loss function to compare the embeddings. Specifically, we use the KRes model, described above, as the backbone with an added linear layer to achieve a 128-dimensional representation. The projection head consists of two multilayer perceptron (MLP) layers with ReLU activation and produces a 64-dimensional embedding. For the loss function, we employ normalized temperature-scaled cross-entropy loss (NT-Xent), as described in [17]. The loss is applied to the embeddings, obtained after the projection head, but we ultimately utilize the representations for downstream task classification, discarding the projection head after training. This architecture can be seen in Fig. 1.

### C. Training

For training the self-supervised model, we use the 4.2 million ECG from the MGH dataset. For this model, we acquire two views of the same sample by splitting each 10-second ECG into two consecutive non-overlapping 5-second ECG views. These segments were then shuffled before being fed into the model so that the model does not inherently know which segment precedes the other. This shuffling is crucial for the contrastive learning framework as it introduces variability, helping the model learn robust and invariant representations of the ECG data. The contrastive model was trained using these ECG segments until the minimum validation loss was achieved, ensuring optimal learning of the ECG representations. This optimization, ideally, closely aligns the representations of the 5-second segments from the same 10-second ECG in the latent space, while driving other samples away. Once the training for this contrastive model is completed, we conduct linear probing and fine-tuning experiments for the task of interest.

We utilize the labeled PTB-XL dataset for multiple purposes: linear probing and fine-tuning the contrastive model, as well as training the supervised model. To do so, the PTB-XL dataset is split in a stratified manner into training-validation-holdout sets, with 10600 ECG in the training set and 1320 in the holdout set. We randomly select either the first or second 5-second section from the 10-second 12-lead ECG for the supervised, fine-tuning, and linear probing training.

In the supervised setup, we train the supervised model with an input of a 5-second 12-lead ECG to a classification prediction and a corresponding binary label (1 = presence of CD, 0 = normal) with binary cross-entropy loss until a minimum validation loss is achieved. We conducted a hyperparameter search over the learning rate for optimal results.

For linear probing, the weights of the model were frozen and the model was then used to obtain 128-dimensional representations of the data in the PTB-XL dataset. A logistic regression model was trained on these representations to classify positive and negative samples, similar to the supervised approach. We optimized the learning rate and temperature hyperparameters to achieve the best performance.

For fine-tuning, the pretrained weights of the KRes backbone from the best-performing self-supervised model were utilized as the initialization for the training setup. All weights were left unfrozen, allowing the entire network to adapt during fine-tuning. This approach allowed the model to leverage the rich, pre-learned features while also tailoring specifically for the supervised task and the new dataset. Additionally, we conducted a hyperparameter sweep to optimize the learning rate.

All models were implemented using Pytorch and Pytorch Lightning, and hyperparameter sweeps were conducted using Hydra. We used the same training, validation, and holdout splits of the PTB-XL dataset to ensure a clear comparison between all methods: 80% of the data for training, 10% for validation, and 10% for holdout.

For supervised training, the best performance was achieved with a learning rate of 0.002. In contrast, self-supervised training benefited most from a lower learning rate of 0.0001 and a temperature of 0.23. For fine-tuning on the pretrained backbone, we found a learning rate of 0.0009 was optimal.

The discriminatory ability of the three modeling approaches is evaluated with the Area Under the receiver-operating Curve (AUC), sensitivity, specificity, and the predictive values on the holdout test set from the PTB-XL dataset.

## IV. Results

Comparative analysis of classification tasks for the three learning approaches is presented in Table I. Given the imbalanced distribution of the two classes in the dataset, AUC represents a balanced performance comparison among the methods. There is no significant difference among the AUC values for these methods, which is an interesting fact when we analyze the other metrics. Observation of the sensitivities and the specificities exposes the fact that the supervised and the linear probing express balanced sensitivity and specificity, around 90%, which is similar to the criteria used on the training set to acquire the decision thresholds. But the fine-tuning method fails to generalize to the holdout set, the sensitivity drops to about 80% for the threshold that achieved 90% sensitivity on the training set. Similarly, the positive and negative predictive values represent the post-test probabilities of the models for the threshold chosen with the 35% prevalence.

**TABLE I.** Performance Comparisons Among Three Learning Approaches.

| Models | AUC<sup>a</sup> [95% CI] | Sensitivity [95% CI] | Specificity [95% CI] | PPV<sup>b</sup> [95% CI] | NPV<sup>b</sup> [95% CI] |
| --- | --- | --- | --- | --- | --- |
| **Supervised** | 96.35 [95.96,96.74] | 88.12 [84.82,90.92] | 91.00 [88.89,92.84] | 84.12 [81.03,86.80] | 93.41 [91.70,94.78] |
| **Self-Supervised + Linear Probing** | 96.00 [95.57,96.44] | 90.06 [86.97,92.63] | 87.73 [85.35,89.86] | 79.89 [76.81,82.65] | 94.23 [92.54,95.56] |
| **Self-Supervised + Fine-tuning** | 95.56 [95.12,96.02] | 81.64 [77.81,85.06] | 95.56 [93.96,96.84] | 90.87 [87.91,93.16] | 90.59 [88.81,92.11] |

<sup>a</sup>Bootstrapping was used to determine 95% CI. Data was sampled with replacement for 100 iterations.
<sup>b</sup>These values are dependent on disease prevalence, which was 35.1%.

To further explore the generalizability and robustness of these methods, we performed additional experiments limiting the amount of training data available while keeping the number of samples in the holdout test set the same. The performances are shown in Fig 2. Here, we notice that the AUC is relatively unchanged for the self-supervised with linear probing method, while both the supervised and self-supervised with fine-tuning methods severely deteriorate as the number of training samples decreases. This analysis is useful in identifying which approach is better suited for data-scarce scenarios, a common challenge in clinical settings. By comparing the performance of the models with restricted training data, we aimed to highlight the advantages of self-supervised learning methods, particularly linear probing, in leveraging large-scale datasets for improved ECG classification in diverse and resourceconstrained environments.

**Fig. 2.**
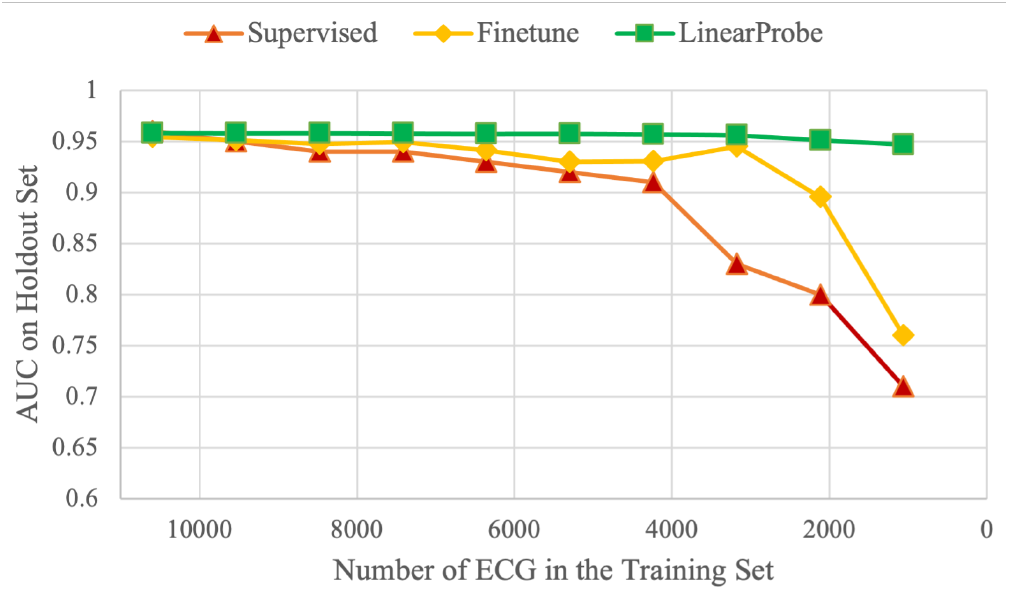
Comparison of learning paradigms across varying training set sizes.

## V. Conclusion

This work demonstrates that self-supervised methodologies can perform on par with supervised models in a data-sufficient setting, but can also perform robustly in data-scarce scenarios. In many healthcare applications, datasets suffer from imbalanced and scant labels. Our results suggest that by leveraging large-scale unlabeled datasets for pretraining, self-supervised methods can provide practical and efficient solutions for enhancing the diagnostic capabilities of deep learning models in cardiac healthcare. We plan to expand this work toward other cardiac abnormalities and to further test the proposed models on more available datasets.

## Data Availability

All data are publicly available online at https://physionet.org/content/ptb-xl/1.0.3/

